# How new openings sustain the income gradient in unhealthy retail: evidence from a statewide establishment panel, Rhode Island, 2016-2025

**DOI:** 10.64898/2026.08.20.26360917

**Authors:** Sai Venkat Mandalapu, Sage Lefebvre, Erica D. Walker

**Author notes:** Corresponding author Erica D. Walker Department of Epidemiology, Brown University School of Public Health, 121 South Main Street, Providence, Rhode Island, 02903, United States of America. Author contributions (CRediT) E.D.W.: Conceptualization, Data curation, Funding Acquisition, Project administration, Resources, Software, Supervision, Validation, Writing – reviewing & editing; **S.V.M.:** Conceptualization, Data curation, Formal analysis, Investigation, Methodology, Software, Validation, Visualization, Writing – original draft, Writing – reviewing & editing; **S.L.:** Validation, Writing – reviewing & editing. Declaration of competing interests The authors declare that they have no known competing financial interests or personal relationships that could have appeared to influence the work reported in this paper.

## Abstract

Unhealthy retail outlets, including liquor stores, bars, convenience stores, and fast food, are concentrated in lower-income neighbourhoods. This is a well-documented cross-sectional fact; the process that sustains it is not. A neighbourhood can hold more because more open there or because those already there survive longer, and these point to different responses. We assembled an establishment-level panel of every business in Rhode Island from 2016 to 2025 (480,923 geocoded establishment-years across nine annual cross-sections), following the same outlets year to year, and classified and counted unhealthy outlets by census tract. We estimated the tract income gradient three ways (negative binomial regression, a concentration index, and a Bayesian spatial model), tested its stability, and decomposed it into openings and closures. The gradient was strong, stable, and robust: about 30 percent fewer unhealthy outlets per resident per standard deviation of higher income, with racial composition and poverty no longer associated once income was included. It was reproduced through entry, not survival: closures were even-handed across income, while new unhealthy outlets opened about 2.2 times as often per resident in the lowest-income tracts as in the highest. This entry was not unhealthy-specific: new healthy food retail tilted toward lower-income tracts at least as strongly, and the unhealthy share of openings did not rise as income fell. The standing burden was nonetheless dominated by convenience stores and off-premise alcohol. Efforts to reshape the retail environment will have more leverage on new openings than on the existing stock, through instruments defined by outlet type.

## 1. Introduction

Neighbourhood retail environments shape what residents consume and, through it, their health. At the neighbourhood level a higher density of alcohol outlets is associated not only with heavier drinking but with injury, crime, and violence (Campbell et al., 2009). The balance of outlets selling energy-dense, nutrient-poor food relative to healthier options, sometimes called a food swamp, predicts obesity rates better than the simple absence of supermarkets (Cooksey-Stowers et al., 2017), and the local food environment is associated, if inconsistently, with diet (Caspi et al., 2012). A common thread across these literatures is that what matters is not only the absence of healthy options but the presence and relative density of unhealthy ones, and that this presence tends to cluster in the same disadvantaged neighbourhoods that carry other social and environmental burdens.

These food and alcohol outlets are not evenly distributed. Lower-income neighbourhoods, and in the United States often neighbourhoods with larger Black and Hispanic populations, tend to have fewer supermarkets and a higher relative density of limited-assortment grocery, convenience, and liquor outlets (Moore and Diez Roux, 2006). The disparity has been replicated across settings and outlet types and is one of the more durable findings in the field. It is almost always established the same way, from a single year of outlet locations regressed on the sociodemographic composition of an area.

What is far less understood is how this distribution arises and why it persists. A snapshot can show that lower-income neighbourhoods hold more unhealthy outlets, but it cannot show how they came to. Two distinct processes generate the standing distribution at any moment. New outlets open in some places more than others, and existing outlets close in some places more than others. A neighbourhood can accumulate unhealthy outlets because more of them open there, because the ones there survive longer, or both. These processes have different drivers and, importantly, different points of leverage for anyone who wants to change the pattern.

Cross-sectional data cannot separate them, and even longitudinal studies of the food environment have usually relied on short follow-up or modest samples and have rarely modelled openings and closures as distinct flows (Lovasi et al., 2023). Separating them requires following individual establishments over time.

In this paper we assemble a comprehensive establishment-level panel for the entire state of Rhode Island from 2016 to 2025, in which the same businesses are tracked from one year to the next. This lets us move the question from where unhealthy outlets are to how they get there. We first confirm the standing income gradient in the unhealthy retail environment, that is, the tendency for such outlets to be more densely concentrated per resident in lower-income tracts (we use ‘concentration’ and ‘income gradient’ interchangeably for this pattern), and show that it is stable across the decade and robust to model choice and spatial structure. We then decompose it into its two generating flows, openings and closures, to ask which one carries the gradient. We treat neighbourhood income as the primary axis throughout, and report racial composition and poverty as correlates that may or may not survive adjustment for income. Finally, as a secondary analysis, we ask whether a measure of historical disinvestment, the Historic Redlining Indicator, adds anything beyond present-day income. Distinguishing openings from survival is not merely descriptive: the two processes imply different points of intervention, so identifying which one sustains the gradient tells local licensing and zoning authorities, and researchers characterising neighbourhood health environments, where any leverage actually lies. The contribution is to replace a static description with a mechanism, using a complete statewide census of establishments rather than a sample.

## 2. Methods

### 2.1 Setting and business data

The study covers the entire state of Rhode Island. We used annual business establishment files for Rhode Island from Data Axle USA (formerly InfoUSA), one file per year from 2016 through 2025. Each record lists a business name, a primary North American Industry Classification System (NAICS) code and a Standard Industrial Classification (SIC) code, geographic coordinates, and a persistent establishment identifier. The validity of commercial business lists for characterising the retail food environment has been assessed directly, including for the InfoUSA and Data Axle lineage, and these databases tend to be reasonably complete and well geocoded but to carry classification error that is heaviest for restaurants (Liese et al., 2010; Han et al., 2012). We return to these limitations in the Discussion.

The ten annual files spanned 2016 to 2025. The 2019 file duplicated the records of an adjacent year and was removed, leaving nine annual cross-sections (Supplementary Table S1) and 480,923 geocoded establishment-years in total (Table 1). We confirmed that the establishment identifier was stable across files, linking the same business from one year to the next, so the nine cross-sections constitute an establishment-level panel rather than a series of independent snapshots. This panel structure is what lets us separate the standing distribution of outlets from the openings and closures that generate it.

**Table 1.**
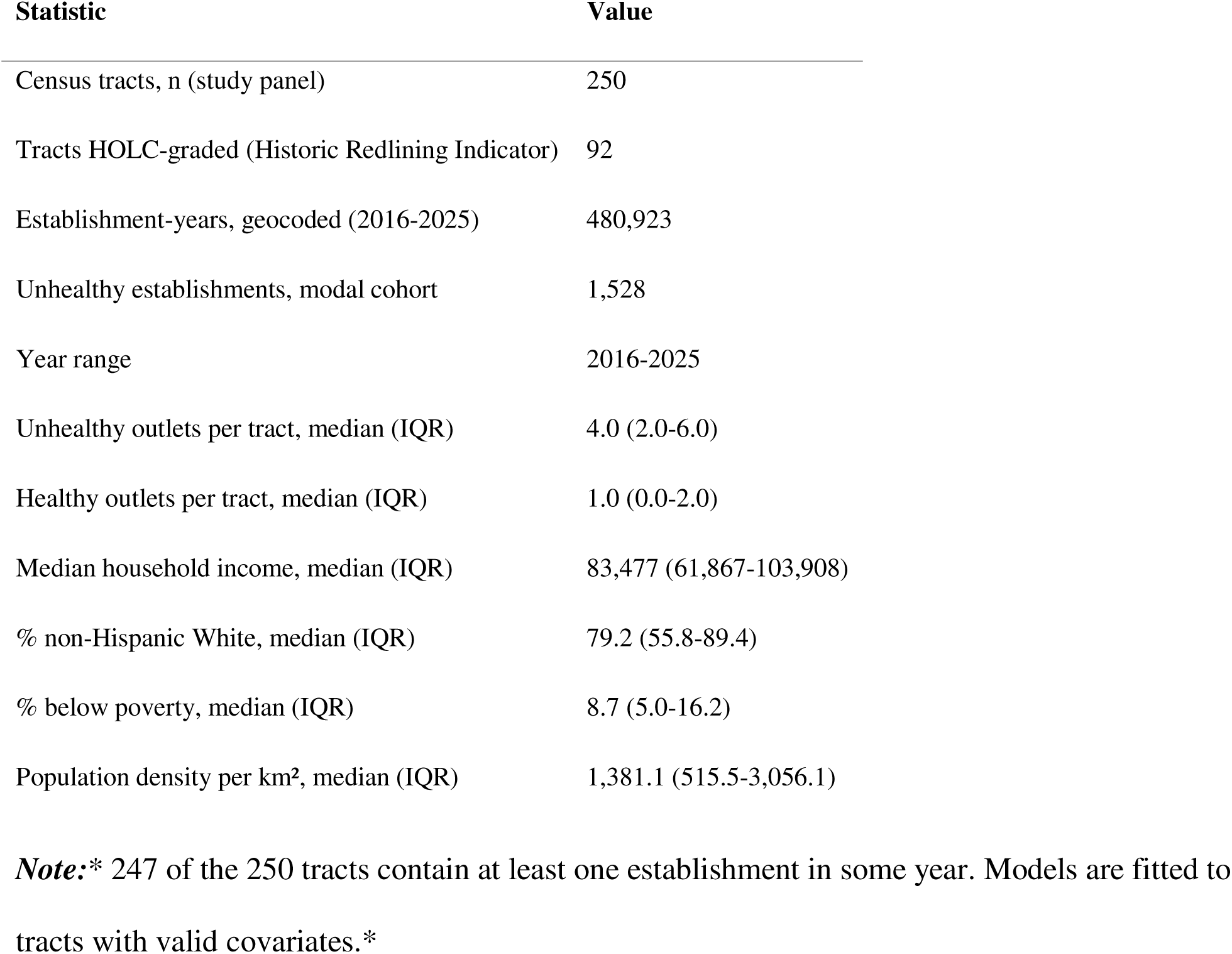
Study panel, most recent year unless noted.

Rhode Island is well suited to this question. It is compact enough that a single commercial database can cover every establishment in the state, so the openings and closures that generate the standing distribution can be observed directly rather than estimated from a sample, yet it contains substantial socioeconomic heterogeneity across neighbourhoods, from the lower-income urban cores of Providence, Pawtucket, Woonsocket, and Central Falls to affluent suburban and shoreline towns. The state also has a distinctive alcohol-retail structure. Under its three-tier system of alcohol regulation, grocery and convenience stores in Rhode Island cannot hold off-premise retail liquor licences, so beer, wine, and spirits are sold almost entirely through dedicated liquor stores rather than within general retailers as they are in many other states (Rhode Island Department of Business Regulation, 2023). This makes off-premise alcohol a cleanly bounded, separately licensed outlet type here, which sharpens the interpretation of the alcohol component of the unhealthy retail environment.

### 2.2 Classification of establishments

We classified each establishment from its industry codes rather than its name, treating NAICS as authoritative and using SIC to resolve missing or ambiguous NAICS values. NAICS revised two relevant codes during the study period, which we reconciled across the 2022 to 2023 boundary so that each category is consistent over time: liquor stores from 445310 to 445320, and convenience stores from 445120 to 445131.

We defined the unhealthy retail environment as four establishment types: liquor stores, drinking places (bars and taverns), convenience stores, and fast food. Because the NAICS eating-place codes do not separate fast food from full-service restaurants, fast food was identified within those codes using a curated list of national and regional quick-service chains. As a healthy comparison set we used supermarkets and grocery stores together with produce markets. Coffee, donut, and ice cream outlets were treated as neutral and counted in neither set. Tobacco-only retailers were excluded.

To check the classification we drew a stratified random sample of 360 establishment-records, 40 in each of the nine assigned categories, and reviewed each one by hand against its business name and its full NAICS and SIC descriptions. Overall agreement with the assigned category was 95.0% (342 of 360). Per-category agreement ranged from 90% to 100% and was 92.5% or higher for every unhealthy category (convenience 92.5%, drinking places 97.5%, fast food 100%, liquor stores 97.5%; Supplementary Table S8). Most disagreements were records that carried a food- retail industry code in the source data for a business that was not a food outlet, such as a car wash or a nonprofit coded as a grocery, rather than errors of category logic.

### 2.3 Geographic and demographic data

We assigned each geocoded establishment to a 2020 census tract using full-resolution TIGER/Line tract boundaries. A small number of coastal establishments fell just outside the land polygons because of geocoding offset; points lying outside all tracts were assigned to the nearest tract within 2 km, and the few points falling outside Rhode Island were dropped. The panel contains 250 tracts, of which 247 hold at least one establishment in some year.

Tract socioeconomic and demographic measures came from the American Community Survey 2018 to 2022 five-year estimates: median household income, the percentage of residents who are non-Hispanic White, the percentage of residents below the federal poverty line, total population, and land area, from which we computed population density per km².

### 2.4 Historic redlining

To represent the legacy of historical disinvestment we used the Historic Redlining Indicator (Meier and Mitchell, 2023), which overlays the 1930s Home Owners’ Loan Corporation residential security grades onto modern census tracts and summarises them as a continuous score, with higher values denoting more severe historical redlining. Ninety-two Rhode Island tracts received a grade and could be linked by tract identifier.

### 2.5 Exposure and outcome measures

The primary measure of the unhealthy retail environment was the count of unhealthy outlets in a tract, modelled as a rate relative to resident population through a log population offset. As a secondary specification we modelled the count relative to land area through a log land-area offset, which describes physical density rather than per-person availability.

### 2.6 Statistical analysis

We standardised all continuous tract covariates (income, percentage non-Hispanic White, poverty rate, log population density, and the redlining indicator) to mean zero and unit standard deviation, so that estimates are interpretable per standard deviation and comparable across covariates.

#### Cross-sectional distribution

We modelled tract unhealthy outlet counts with negative binomial regression, entering income, percentage non-Hispanic White, poverty, and log population density first singly and then jointly, under both the per-capita offset and the per-area offset (Table 2). Income was the primary socioeconomic axis.

**Table 2.**
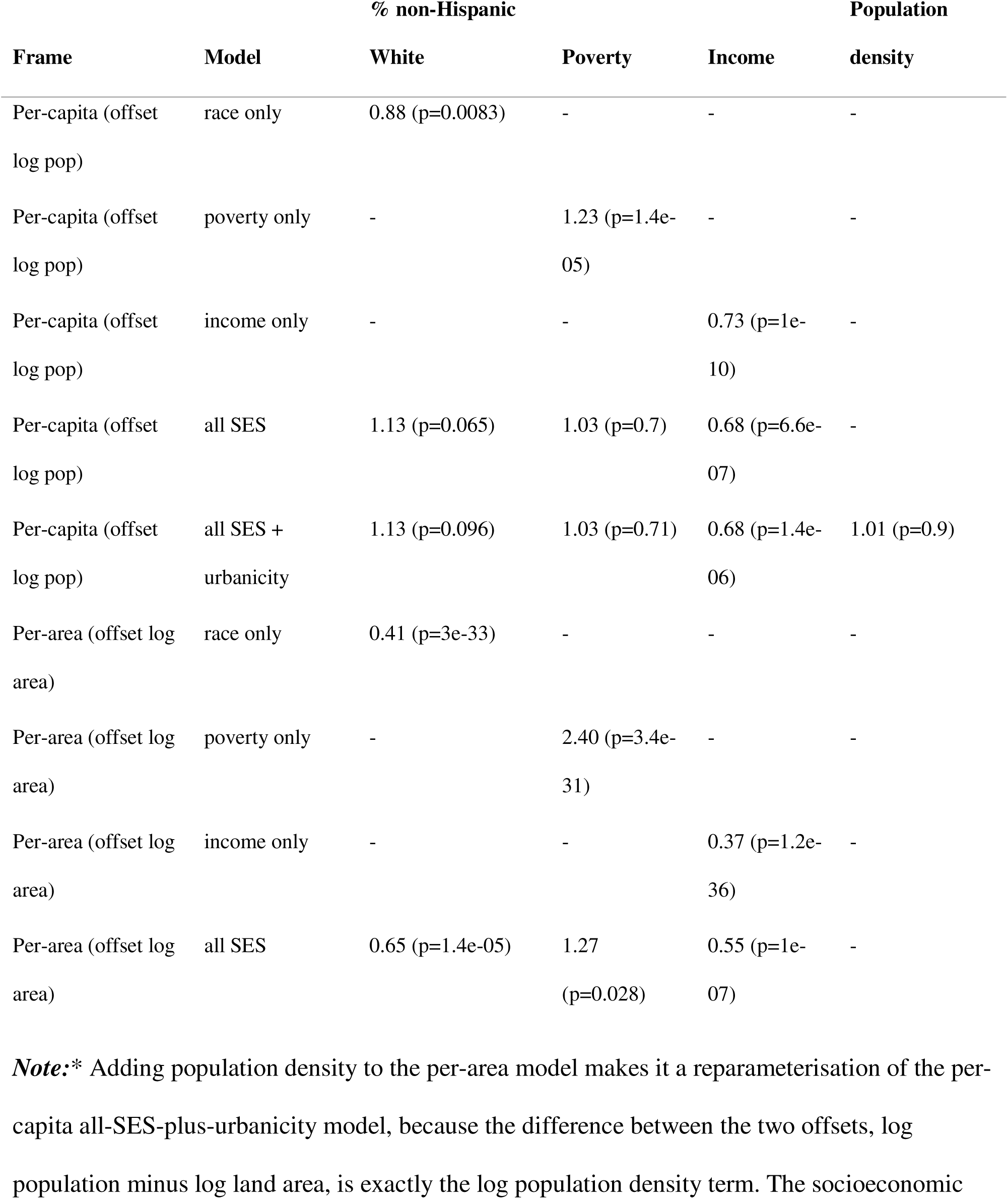
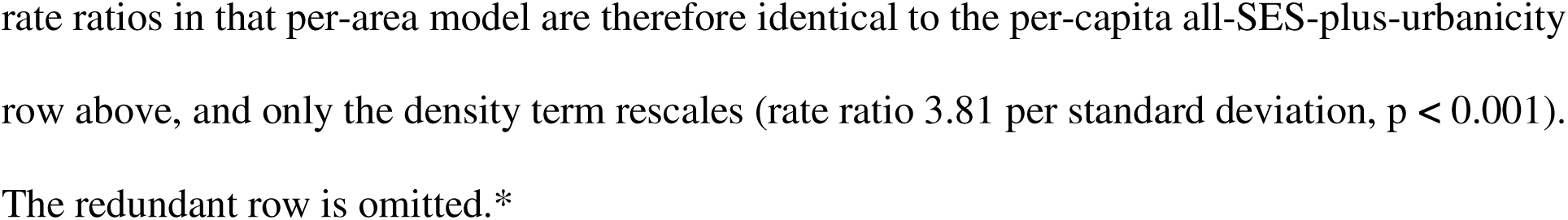
Cross-sectional negative binomial models of unhealthy outlet counts, most recent year. Cells are rate ratios per standard deviation (p value).

| Frame | Model | % non-Hispanic |  |  | Population density |
| --- | --- | --- | --- | --- | --- |
|  |  | White | Poverty | Income |  |
| Per-capita (offset log pop) | race only | 0.88 (p=0.0083) | - | - | - |
| Per-capita (offset log pop) | poverty only | - | 1.23 (p=1.4e-05) | - | - |
| Per-capita (offset log pop) | income only | - | - | 0.73 (p=1e-10) | - |
| Per-capita (offset log pop) | all SES | 1.13 (p=0.065) | 1.03 (p=0.7) | 0.68 (p=6.6e-07) | - |
| Per-capita (offset log pop) | all SES + urbanicity | 1.13 (p=0.096) | 1.03 (p=0.71) | 0.68 (p=1.4e-06) | 1.01 (p=0.9) |
| Per-area (offset log area) | race only | 0.41 (p=3e-33) | - | - | - |
| Per-area (offset log area) | poverty only | - | 2.40 (p=3.4e-31) | - | - |
| Per-area (offset log area) | income only | - | - | 0.37 (p=1.2e-36) | - |
| Per-area (offset log area) | all SES | 0.65 (p=1.4e-05) | 1.27 (p=0.028) | 0.55 (p=1e-07) | - |

**Note:\*** Adding population density to the per-area model makes it a reparameterisation of the per-capita all-SES-plus-urbanicity model, because the difference between the two offsets, log population minus log land area, is exactly the log population density term. The socioeconomic
rate ratios in that per-area model are therefore identical to the per-capita all-SES-plus-urbanicity row above, and only the density term rescales (rate ratio 3.81 per standard deviation, $p < 0.001$ ). The redundant row is omitted.\*

#### The income gradient, triangulated

Because a single model can be sensitive to its specification, we estimated the income gradient three ways (Table 3). First, a negative binomial incidence rate ratio for unhealthy outlets per standard deviation of income. Second, a population-weighted health concentration index that ranks tracts by income (Wagstaff et al., 1991), with confidence intervals from a nonparametric bootstrap. Third, a Bayesian spatial model: the BYM2 parameterisation of the Besag-York-Mollié model (Besag et al., 1991; Riebler et al., 2016), specified as a Poisson rate model with population as the expected count, income and log density as fixed effects, and a combined spatially structured and unstructured tract random effect on a queen-contiguity adjacency, with penalised-complexity priors and a scaled spatial component. We fitted it using the integrated nested Laplace approximation (Rue et al., 2009). This model also returns phi, the proportion of the random-effect variance that is spatially structured; a low value indicates little residual spatial clustering once the covariates are accounted for.

**Table 3.** The income gradient estimated three ways.

| Method | Estimate | 95% interval |
| --- | --- | --- |
| Negative binomial, per-capita rate ratio per SD income | 0.704 | 0.621 to 0.798 |
| Concentration index by income, most recent year | -0.1711 | -0.2207 to -0.1143 |
| BYM2 Bayesian spatial, rate ratio per SD income ($\phi = 0.061$) | 0.690 | 0.609 to 0.781 |

#### Stability over the decade

We refitted the per-capita income model separately in each of the nine years to test whether the gradient was stable over time (Supplementary Table S3), and computed the concentration index for income and for the non-Hispanic White share in every year (Supplementary Table S2).

#### Openings and closures

To ask how the standing distribution is produced rather than only describing it, we used the panel to model two flows among the cohort of 1,528 establishments whose modal category across their observed lifespan was unhealthy. We defined an opening, which we also refer to as entry, as the first year in which a persistent establishment identifier appeared anywhere in the data, in any industry category, and a closure as an establishment’s last observed year falling before the end of the panel. Defining openings at the level of the establishment identifier rather than the category is deliberate: a business that was present in an early year but recorded under a different code, and only later classified as unhealthy, is not counted as a new opening, because its identifier already existed. Of the 1,528 establishments, 447 first appeared after 2016 (openings) and 459 were last seen before 2025 (closures), with 1,069 still present in the final year.

For closures we fitted a discrete-time survival model (Allison, 1982): each establishment contributed one record per year from its first to its last observed year, the event was closure, the link was complementary log-log, year indicators carried the baseline hazard, and income and log density entered as covariates. For openings we modelled the count of newly appearing unhealthy establishments per tract-year with negative binomial regression, a log population offset, and year indicators. In both flow models we clustered standard errors by tract (Cameron and Miller, 2015). We also summarised the two flows descriptively, as the closure rate and the number of openings per 1,000 residents across income tertiles (Table 5).

**Table 4.** Dynamics models: closure hazard and openings. Estimates per standard deviation.

| Analysis | Sample | Term | Estimate | Lo | Hi | p |
| --- | --- | --- | --- | --- | --- | --- |
| Closure hazard | full state | income | 1.105 | 0.986 | 1.239 | 0.0861 |
| Closure hazard | full state | log pop density | 1.206 | 1.072 | 1.356 | 0.00185 |
| Closure hazard | HOLC subset | redlining (HRI) | 1.172 | 1.005 | 1.367 | 0.0428 |
| Closure hazard | HOLC subset | income | 1.134 | 0.971 | 1.325 | 0.111 |
| Closure hazard | HOLC subset | log pop density | 1.25 | 1.073 | 1.457 | 0.00423 |
| Openings (per-capita) | full state | income | 0.734 | 0.595 | 0.906 | 0.00392 |
| Openings (per-capita) | full state | log pop density | 1.064 | 0.89 | 1.273 | 0.496 |
| Openings (per-capita) | HOLC subset | redlining (HRI) | 1.303 | 0.954 | 1.78 | 0.0955 |
| Openings (per-capita) | HOLC subset | income | 0.804 | 0.583 | 1.11 | 0.185 |
| Openings (per-capita) | HOLC subset | log pop density | 0.878 | 0.706 | 1.093 | 0.246 |

**Table 5.** Establishment flows by tract income tertile: the per-establishment closure rate and, per 1,000 residents, closures and openings. Tertiles are tract-level (equal-count thirds of tracts by median household income); rates are aggregate (events divided by tertile population).

| Income tertile | Closure rate (per establishment) | Closures per 1,000 | Openings per 1,000 |
| --- | --- | --- | --- |
| Low | 0.289 | 0.540 | 0.586 |
| Middle | 0.307 | 0.441 | 0.398 |
| High | 0.314 | 0.294 | 0.261 |

Two features of the panel required handling. First, the 2016 and 2017 files differ in classification composition in a way that looks like a one-time coverage or coding shift rather than real change on the ground (Supplementary Table S1). Our primary analysis uses all nine cross-sections; as a sensitivity analysis we repeated both flow models on a 2018-onward window, treating 2016 and 2017 as a settling-in period (Supplementary Table S7). Second, because the 2019 file was dropped, person-periods are defined over observed waves, with 2018 and 2020 treated as adjacent; 68 cohort establishments are last seen in 2018 and coded as closed, and since 2019 is unobserved a 2018 closure cannot be distinguished from a 2019 one (Supplementary Table S6).

#### Specificity and composition of openings

To test whether the openings gradient was specific to unhealthy retail rather than a feature of commercial entry generally, we counted openings for two comparison sets alongside the unhealthy set, using the same establishment-identifier definition of an opening (first appearance after 2016) and the same per-tract-year negative binomial model with a log population offset and year indicators: all openings, defined as the first appearance of any establishment in any category, and healthy-retail openings, restricted to supermarkets, grocery stores, and produce markets. We then modelled the unhealthy composition of entry directly in two ways. First, a fractional logit (quasibinomial) model of the count of unhealthy openings out of all openings in a tract-year on standardised income and log density, which asks whether the unhealthy share of new openings changes with income. Second, a negative binomial for unhealthy openings with the log of total openings as an offset, which expresses unhealthy openings relative to total openings on the rate-ratio scale. We also refitted the openings model with a BYM2 spatial random effect (Supplementary Material S5), reported category-specific cross-sectional and openings gradients and re-estimated the flows excluding bars (Supplementary Table S10), and summarised net flow and per-capita stock by income tertile (Supplementary Table S9). Standard errors in the count models were clustered by tract.

#### Secondary analyses with redlining

Within the subset of HOLC-graded tracts we added the standardised Historic Redlining Indicator to the cross-sectional, by-year, and flow models (Supplementary Tables S3, S4, and Table 4). These are reported as a secondary analysis. Analyses were carried out in R. Spatial operations used the sf package, and the Bayesian spatial model used R-INLA.

## 3. Results

### 3.1 The retail landscape

Across the nine cross-sections the panel contained 480,923 geocoded establishment-years in 250 tracts (Table 1). In the most recent year a typical tract held a median of 4 unhealthy outlets (interquartile range 2 to 6) and 1 healthy outlet (0 to 2). Tracts varied widely on the measures we use as predictors: median household income ran from about 62,000 at the 25th percentile to about 104,000 at the 75th, the non-Hispanic White share from 56 to 89 percent, and the poverty rate from 5 to 16 percent (Table 1).

At the aggregate level the unhealthy retail environment was close to stable across the decade. Summing the four unhealthy categories, the statewide count was 1,064 in 2016 and 1,122 in 2025, with convenience stores increasing and fast food declining slightly over the period (Supplementary Table S1). The one conspicuous exception was a single-year jump between 2016 and 2017, when recorded convenience stores rose by 56 percent and bars fell by 55 percent; as we show below, this was a coverage and classification shift between the two earliest files rather than real change on the ground, and it did not affect the openings analysis (Section 3.5; Supplementary Tables S1, S6). Healthy outlets were also roughly stable, rising from 271 to 317, mostly through supermarkets and grocery stores. The total number of unhealthy outlets was therefore not the story; their distribution was (Figure 1).

**Figure 1.**
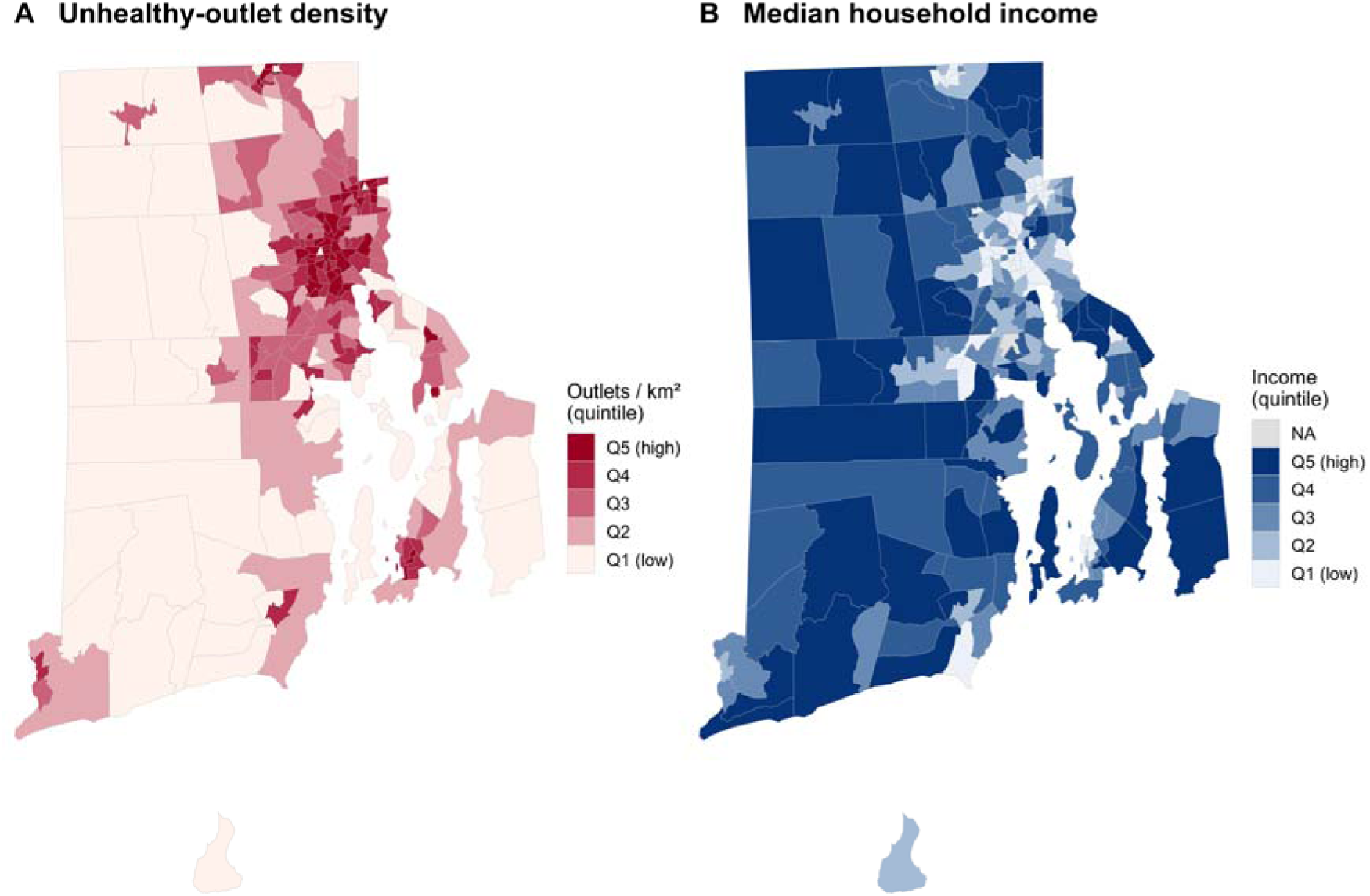
Unhealthy-outlet density and median household income across Rhode Island census tracts, most recent year (2025). (A) Unhealthy outlets per km², in quintiles. (B) Median household income, in quintiles. The tracts with the densest unhealthy retail environments correspond closely to the lower-income tracts. Tracts without an estimate are shown in grey. Boundaries are 2020 TIGER/Line cartographic tract boundaries.

### 3.2 The gradient tracks income, not race or poverty

In per-capita models, higher-income tracts had fewer unhealthy outlets, and income was the axis that survived adjustment (Table 2). Entered on its own, each socioeconomic measure was associated with the count: the income rate ratio was 0.73 per standard deviation (p < 0.001), the poverty rate ratio 1.23 (p < 0.001), and the non-Hispanic White rate ratio 0.88 (p = 0.008). When income, race, poverty, and population density were entered together, only income remained: its rate ratio was 0.68 per standard deviation (p < 0.001), while the non-Hispanic White share (1.13, p = 0.096) and poverty (1.03, p = 0.71) were no longer associated. The per-area specification told the same story about income once density was included, with physical density itself the dominant determinant of how many outlets occupy a given area (rate ratio 3.81 per standard deviation, p < 0.001; Table 2). In short, the racial and poverty associations seen in simpler models reflected income, not something separate from it.

### 3.3 The income gradient is robust

Three different methods agreed on the size of the income gradient (Table 3). The negative binomial rate ratio was 0.704 per standard deviation of income (95% confidence interval 0.621 to 0.798), about 30 percent fewer unhealthy outlets per resident for each standard deviation of higher income. The population-weighted concentration index for the most recent year was -0.171 (95% confidence interval -0.221 to -0.114), confirming that unhealthy outlets fall disproportionately in lower-income tracts. The Bayesian spatial model returned a rate ratio of 0.690 (95% credible interval 0.609 to 0.781), essentially unchanged after allowing for spatial correlation between neighbouring tracts. The spatial share of residual variation was low (phi = 0.061), pointing to little structured residual clustering beyond what income and density already capture. Given the penalised-complexity prior placed on this parameter, which favours smaller structured shares, we read this as evidence against strong leftover spatial structure rather than as a precise variance decomposition, and we do not claim that the covariates fully absorb the spatial pattern. The gradient was not an artefact of one model’s functional form or of unmodelled geography (Figure 2).

**Figure 2.**
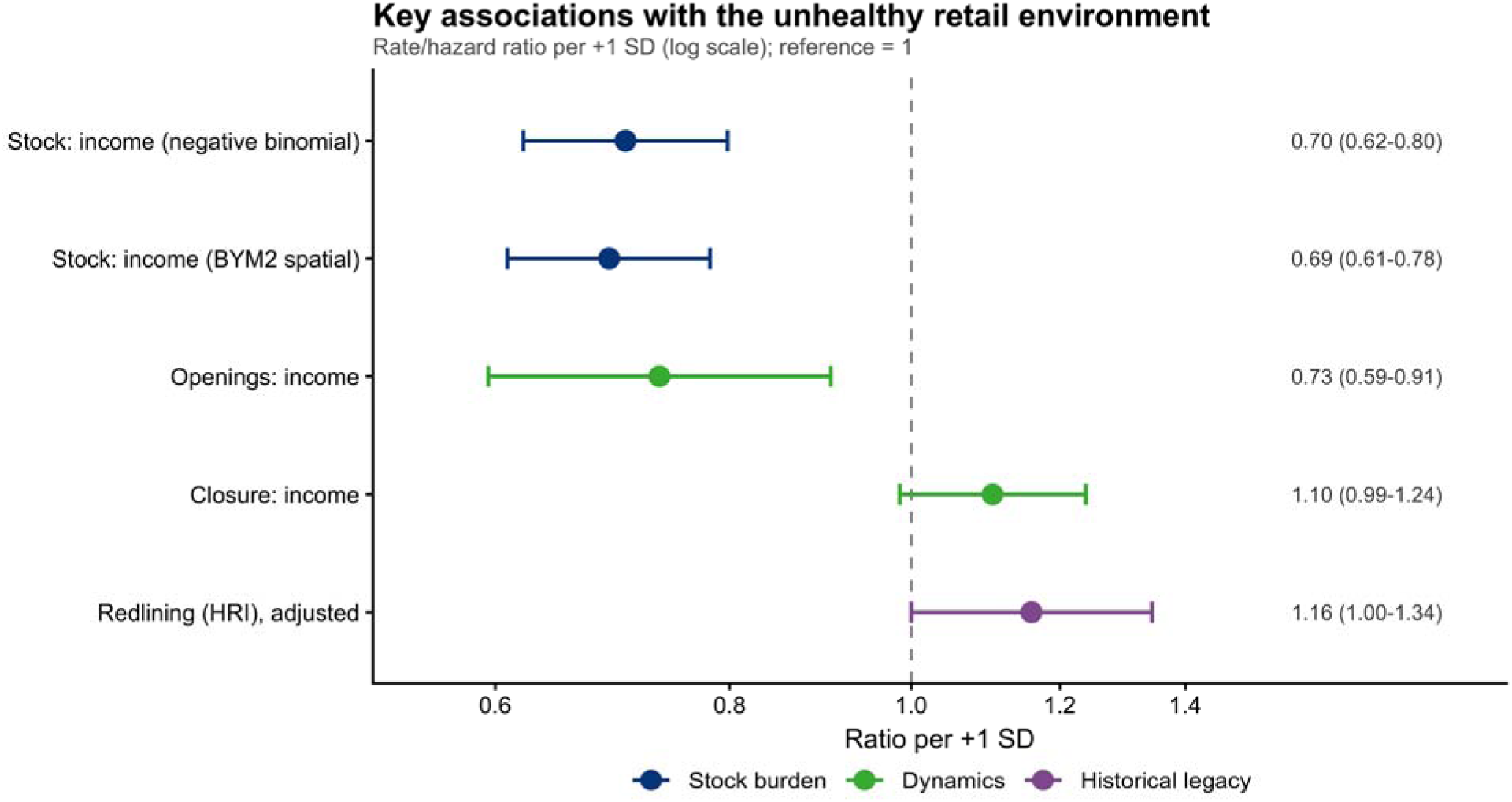
Key associations with the unhealthy retail environment, per one standard deviation (log scale; reference = 1). Stock burden: the cross-sectional income rate ratio from the negative binomial model (0.70, 95% confidence interval 0.62 to 0.80) and from the BYM2 spatial model (0.69, 0.61 to 0.78). Dynamics: the income rate ratio for openings (0.73, 0.60 to 0.91) and the income hazard ratio for closures (1.11, 0.99 to 1.24), both full-state. Historical legacy: the Historic Redlining Indicator rate ratio, adjusted for current income and density within HOLC-graded tracts (1.16, 1.00 to 1.34). Horizontal bars are 95% confidence or credible intervals.

### 3.4 The income gradient is stable across the decade

The gradient was a fixed feature of the period rather than a recent development. The per-capita income rate ratio stayed within a narrow band, from about 0.70 to 0.73, in every year from 2016 to 2025 (Supplementary Table S3), and the income concentration index stayed between about - 0.15 and -0.17 across all nine years (Supplementary Table S2, Figure 3). Whatever produced the gradient was operating consistently throughout.

**Figure 3.**
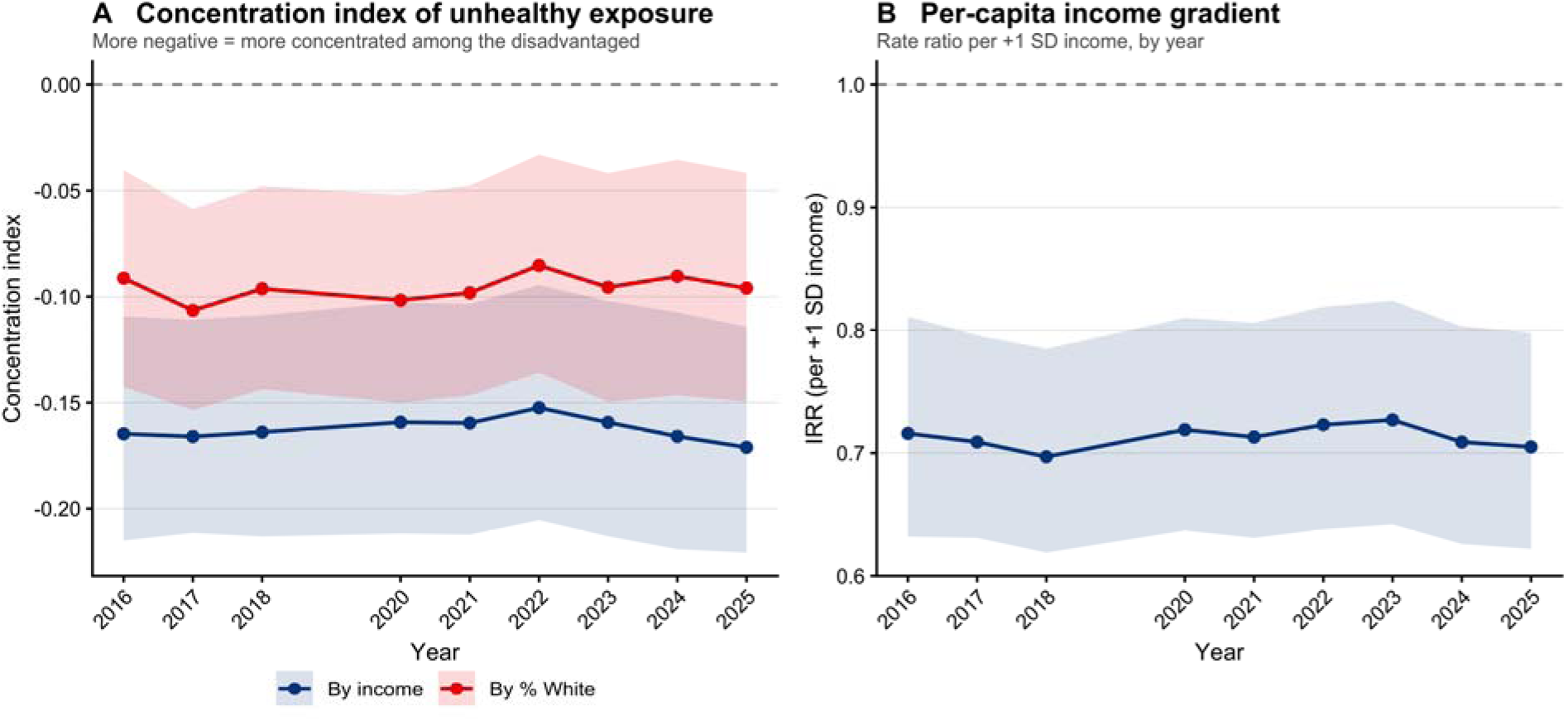
The income gradient was stable across the decade. (A) Population-weighted concentration index of unhealthy-outlet exposure by year, ranking tracts by income (blue) and by non-Hispanic White share (red); more negative values indicate greater concentration in lower- income (or lower-White-share) tracts, and shaded bands are 95% confidence intervals. (B) Per- capita negative binomial income rate ratio for unhealthy outlets by year, per one standard deviation of higher income (reference = 1), with a 95% confidence band. Both quantities remain near-constant from 2016 to 2025; the 2019 wave is absent because the duplicate file was dropped.

### 3.5 The mechanism: openings, not closures

We then asked how a stable standing gradient is generated, by modelling the two flows that can produce it (Table 4, Table 5). Of the 1,528 establishments in the modal-unhealthy cohort, 447 opened during the period and 459 closed.

Closures did not carry the gradient. In the discrete-time survival model, the closure hazard of unhealthy establishments was not associated with tract income (hazard ratio 1.105 per standard deviation, 95% confidence interval 0.986 to 1.239, p = 0.086). Descriptively, the closure rate over the period was nearly flat across tract income tertiles, and if anything slightly higher in higher-income tracts: 0.289 in the lowest-income third of tracts, 0.307 in the middle, and 0.314 in the highest (Table 5). An unhealthy outlet in a lower-income tract was no more likely to close than one in a higher-income tract, and the point estimates run mildly in the opposite direction to the standing gradient. Closures did rise with population density (hazard ratio 1.206, p = 0.002), consistent with higher commercial turnover in denser areas, but this runs alongside the income question rather than answering it.

Openings carried the gradient. New unhealthy establishments opened disproportionately in lower-income tracts. The opening rate fell with income (rate ratio 0.734 per standard deviation, 95% confidence interval 0.595 to 0.906, p = 0.004) and was independent of population density (density rate ratio 1.064, p = 0.50). Descriptively, the lowest-income third of tracts saw about 2.2 times as many openings per 1,000 residents as the highest-income third (0.586 versus 0.261 per 1,000, with the middle tertile at 0.398; Table 5). Placed in the same units, closures were also higher per resident in the lowest-income third (0.540 versus 0.294 per 1,000), because there is more unhealthy stock there to close; the two flows differ not in their rate per establishment, which is nearly flat, but in that openings outpace closures in lower-income tracts and fall short of them in higher-income tracts (Table 5).

Because openings were defined by when an establishment identifier first appeared, the apparent surge in convenience stores between 2016 and 2017 did not enter the opening counts as new entry. Of the cohort, 1,081 establishments were already present in 2016, and only 20 first appeared in 2017, of which 12 were convenience stores (Supplementary Table S6); the 56 percent rise in the convenience count over that year is therefore reclassification of businesses already in operation, not a wave of openings. Restricting both flow models to a 2018-onward window, which excludes the two earliest files entirely, left the result intact: the opening rate fell with income at essentially the same magnitude (rate ratio 0.730 per standard deviation, 95% confidence interval 0.586 to 0.910, p = 0.005), the closure hazard remained unrelated to income (hazard ratio 1.126, p = 0.122), and the descriptive tertile pattern was unchanged, with a similar openings contrast between the lowest- and highest-income thirds (Supplementary Table S7). The closure count is more sensitive than the opening count to where the window starts, but the absence of an income gradient in closures holds either way.

These flows reconcile with the stability of the standing gradient. Over the decade the lowest-income tertile saw more openings than closures (204 versus 188) and its per-capita unhealthy stock edged up, while the middle and highest tertiles ran net negative (140 versus 155 and 103 versus 116); the per-capita stock ratio between the lowest- and highest-income tertiles moved only from about 1.9 to about 2.1 (Supplementary Table S9). A stable gradient is thus not the absence of turnover but its balance: because closures fall at an even rate on a larger low-income stock, they remove more outlets there in absolute terms, offsetting the higher inflow and holding the disparity roughly constant. The openings gradient was also robust to spatial structure; in a BYM2 model with a spatial random effect the opening rate still fell with income (rate ratio 0.739, 95% credible interval 0.622 to 0.879), essentially the non-spatial estimate (Supplementary Material S5).

Taken together, the standing income gradient is reproduced through entry rather than through differential survival. New unhealthy outlets keep arriving more often in lower-income neighbourhoods, while the outlets already present leave at the same rate regardless of neighbourhood income. The pattern is maintained at the front door, not the back (Figure 4).

**Figure 4.**
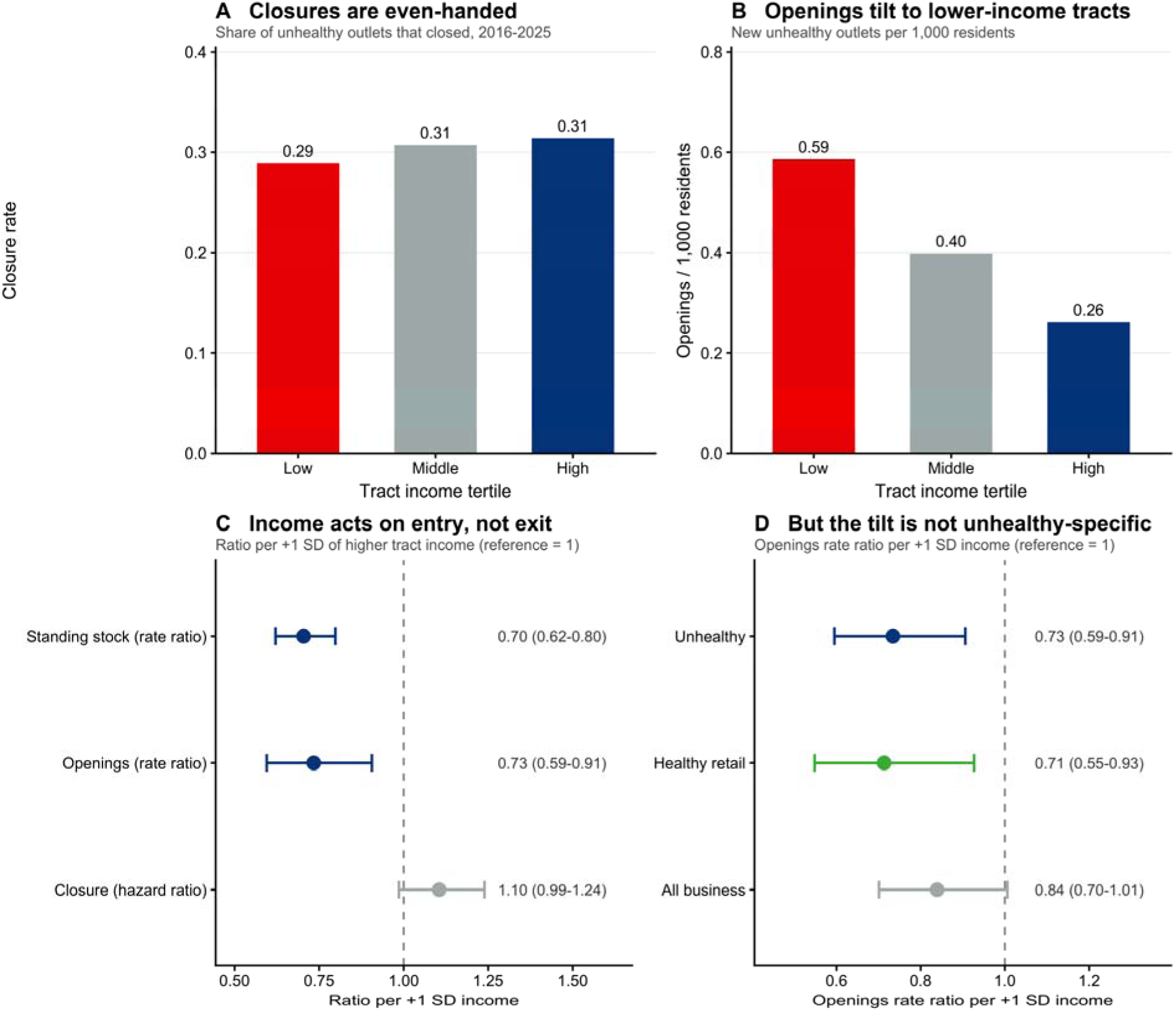
The income gradient in the unhealthy retail environment is produced by entry, not by closure. (A) Share of unhealthy establishments that closed over 2016 to 2025, by tract income tertile; closure rates are nearly flat across tertiles and if anything slightly higher in higher-income tracts (low 0.29, middle 0.31, high 0.31). (B) New unhealthy establishments per 1,000 residents, by tract income tertile; openings are about 2.2 times more common per resident in the lowest-income third than in the highest (0.59 versus 0.26, middle 0.40). (C) Full-state model estimates for tract income, per one standard deviation of higher income (reference = 1): the standing stock of unhealthy outlets falls with income (rate ratio 0.70, 95% confidence interval 0.62 to 0.80) and the opening rate falls with it in near-lockstep (0.73, 0.60 to 0.91), while the closure hazard does not (1.11, 0.99 to 1.24). The standing gradient is reproduced by where unhealthy outlets open, not by which ones survive. (D) Income rate ratio for openings per one standard deviation of higher income (reference = 1), by outlet group: unhealthy 0.73 (0.60 to 0.91), healthy food retail 0.71 (0.55 to 0.93), and all businesses 0.84 (0.70 to 1.01). New openings tilt toward lower-income tracts for unhealthy and healthy retail alike, so the entry that reproduces the unhealthy gradient reflects a broad tilt of commercial entry rather than unhealthy-specific siting.

### 3.6 Entry is not specific to unhealthy retail

The entry that reproduced the gradient was not peculiar to unhealthy outlets. When we counted openings of all kinds, new establishments in general also tended to open more often per resident in lower-income tracts, though this fell just short of significance (rate ratio 0.839 per standard deviation of higher income, 95% confidence interval 0.701 to 1.006), and new healthy food retail tilted toward lower-income tracts at least as strongly as unhealthy retail did (rate ratio 0.713, 0.548 to 0.927). Modelling the unhealthy share of new openings directly, the unhealthy fraction of entry did not rise as income fell (odds ratio 0.933 per standard deviation, 95% confidence interval 0.805 to 1.080, p = 0.35; a specification expressing unhealthy openings relative to total openings gave 0.89, p = 0.076). Lower-income Rhode Island neighbourhoods received more new commercial activity of most kinds, and unhealthy outlets arrived on that broader current rather than being singled out for those neighbourhoods. What sustained the unhealthy gradient was that unhealthy outlets formed part of a commercial inflow that was itself tilted toward lower-income areas, not an unhealthy-specific process of siting. That new healthy retail tilted toward lower- income tracts somewhat more strongly than unhealthy retail (rate ratio 0.713 versus 0.734) raises a question our design does not settle: we do not model the standing distribution of healthy outlets or their closures, so whether this inflow leaves a correspondingly healthy-rich stock in lower- income tracts, or is offset by faster healthy-outlet turnover, is left open.

The standing burden was nonetheless carried by specific formats. All four unhealthy categories showed the same cross-sectional income gradient (rate ratio per standard deviation 0.79 for liquor stores, 0.66 for bars, 0.68 for convenience stores, and 0.68 for fast food, each significant), so pooling them does not mask divergent categories (Supplementary Table S10). The recorded number of bars drifted across the whole panel rather than only at the 2016 to 2017 transition (from 220 in 2016 to 188 in 2025, with a trough near 100 in the intervening years), which makes their year-to-year counts less reliable, and bars accounted for roughly two fifths of the cohort’s raw openings (190 of 447). Even so, excluding bars entirely barely changes the flow result (openings rate ratio 0.757, p = 0.004; closure hazard ratio 1.113, p = 0.11), so the reliance on the noisiest category is not what produces the gradient. Convenience stores and off-premise alcohol, which were coded more reliably and dominated the unhealthy set, therefore carried the concentration on their own (Supplementary Table S10).

### 3.7 Historical redlining (secondary)

Within the 92 HOLC-graded tracts, the Historic Redlining Indicator behaved as a modest correlate of the unhealthy retail environment that largely overlapped with present-day income (Supplementary Table S4). On its own, more severe historical redlining was associated with more unhealthy outlets per resident (rate ratio 1.25 per standard deviation, 95% confidence interval 1.085 to 1.44, p = 0.002). After adjustment for current income and density this association fell to the margin of significance (rate ratio 1.159, 95% confidence interval 1.000 to 1.344, p = 0.050), while income remained clearly associated (rate ratio 0.821, p = 0.014). In the flow models within the graded subset, redlining was associated with a somewhat higher closure hazard (hazard ratio 1.172, p = 0.043) and, more weakly, with more openings (rate ratio 1.303, p = 0.096). Because more severely graded tracts showed both somewhat more closures and somewhat more openings, this read as elevated commercial turnover in those tracts rather than the one-sided entry pattern that carries the income gradient. We interpret these patterns in the Discussion.

## 4. Discussion

In an exhaustive statewide establishment panel, the unhealthy retail environment was consistently concentrated in lower-income Rhode Island neighbourhoods. The concentration was sizeable, about 30 percent fewer unhealthy outlets per resident for each standard deviation of higher income, and it was stable across a decade and robust to model choice and to spatial structure. Neighbourhood racial composition and poverty, which are associated with the unhealthy retail environment in simpler models, were not associated once income was in the model. The new finding is mechanistic. This concentration is reproduced through where new outlets open, not through which outlets survive. Closures were even-handed across the income distribution, if anything marginally more common in higher-income tracts; openings were about 2.2 times more common per resident in the lowest-income tracts than in the highest. The decomposition held when the two earliest annual files were set aside and under a spatial model, so it does not rest on the years most affected by the coverage shift or on unmodelled geography.

A second result qualifies how this entry should be read. The tilt of openings toward lower-income neighbourhoods is not specific to unhealthy retail: new healthy food retail opened disproportionately in lower-income Rhode Island tracts, new businesses more broadly tended to do the same, and the unhealthy share of new openings did not rise as income fell. The unhealthy gradient is therefore reproduced by a broad current of commercial entry running toward lower-income areas, on which unhealthy outlets are carried, rather than by a process that singles those neighbourhoods out for unhealthy formats in particular. This is a weaker claim than unhealthy- specific siting, and an honest one: what our data establish is the flow through which the standing disparity is maintained, not a motive peculiar to unhealthy retail.

This distinction matters because the two processes offer different points of leverage. If the gradient were a survival phenomenon, with unhealthy outlets persisting longer in lower-income areas, attention would turn to what keeps them in business or what crowds out healthier alternatives. Because it is an entry phenomenon, the leverage is on what opens. Our openings result generalises a pattern that has been described for one retail format at a time. The rapid spread of dollar stores, for example, has been concentrated in lower-income and minority neighbourhoods and was identified using models of change rather than snapshots, under the heading of retailer redlining (Shannon, 2021). We find a parallel directional pattern across the full set of unhealthy formats rather than a single one, but with a qualification our single-format predecessors could not see: the tilt of new unhealthy openings toward lower-income neighbourhoods is part of a broader tilt of commercial entry, not an unhealthy-specific one (Section 3.6).

Policies that act on entry, such as licensing limits and zoning, are the natural candidates, but the evidence counsels care about both what they target and what they achieve. The clearest cautionary case is fast-food zoning. South Los Angeles’s 2008 ban on new standalone fast-food restaurants produced no measurable improvement in diet or obesity in the years that followed; if anything, overweight and obesity rose faster in the targeted neighbourhoods than elsewhere in the county (Sturm and Hattori, 2015). The reason is instructive and directly relevant here: the ban reached only standalone fast-food outlets, which were uncommon in the area, while the local unhealthy retail environment was dominated by small food and convenience stores the ordinance did not cover, so only about a tenth of outlets turned over under the rule and their composition did not shift differentially across areas. Our data point to the same hazard, because the unhealthy mix in Rhode Island is dominated by convenience stores and off-premise alcohol more than by standalone fast food, so an entry policy aimed narrowly at one visible format would miss most of what concentrates in lower-income tracts. That policy can bite when it is matched to the dominant format is suggested by the alcohol side: regulating alcohol outlet density through licensing and zoning is an evidence-based strategy for reducing excessive consumption and related harms (Campbell et al., 2009), and in a natural experiment in Atlanta a roughly three percent reduction in on-premise alcohol outlet density tracked a two-fold greater reduction in exposure to violent crime than in comparison areas where density rose (Zhang et al., 2015). Because the inflow into lower-income neighbourhoods is broad and includes the healthier retail those areas also gain, a blunt limit on new outlets would curb desirable openings alongside undesirable ones; the leverage is not in slowing entry as such but in shaping its composition through instruments defined by the outlet types that actually make up the flow. We make no claim that limiting entry would by itself improve health.

The Historic Redlining Indicator behaved as a modest correlate that largely overlapped with present-day income. Its unadjusted association with the unhealthy retail environment is consistent with evidence linking redlining to less healthy food environments (Li and Yuan, 2022) and to present-day health more broadly (Lee et al., 2022), though that comparator concerns food environments specifically while our unhealthy set also includes alcohol outlets, so the comparison is not exact. Once current income and density were included the association fell to borderline significance while income remained robust, and because the redlining score is strongly correlated with the very income and density it predates, we treat it as a historical marker of disinvestment that travels with current income and keep it secondary to the income story.

Set against prior work, the contribution is to supply the missing step. The cross-sectional disparity has been documented many times (Moore and Diez Roux, 2006; Li and Yuan, 2022), and a smaller body of work has modelled change with limited follow-up or samples (Lovasi et al., 2023); by following every establishment in a state across a decade, we show that it is, mechanically, an entry phenomenon.

### 4.1 Implications and future directions

These findings speak to two audiences. For researchers who characterise neighbourhood health environments, the methodological point is that a standing disparity should be read as the net of flows: decomposing it into entry and survival locates the process that sustains it and, with it, the point where change would have to act. For local policymakers, the practical point is that the leverage on the unhealthy retail environment lies in the composition of new openings, addressed through outlet-type licensing and zoning, rather than in the existing stock or in blunt caps on commercial activity. Several extensions follow. The retail environment is better monitored dynamically, from panels that track openings and closures, than from periodic snapshots that cannot tell a growing disparity from a stable one. Our design deliberately kept the healthy side in the background; modelling the standing distribution and turnover of healthy outlets, not only their entry, would show whether the commercial inflow into lower-income neighbourhoods leaves them better or worse supplied over time. And because the entry we document is broadly commercial rather than unhealthy-specific, the open question for both research and policy is why new commercial activity of all kinds concentrates in lower-income neighbourhoods, and whether outlet-type instruments that reshape its composition can be shown to change diet, drinking, or the downstream health outcomes this study did not measure.

### 4.2 Limitations

Several limitations qualify these findings. First, the data are commercial. Business lists such as Data Axle carry classification and completeness error that is heaviest for restaurants (Liese et al., 2010; Han et al., 2012). Our hand validation of a stratified sample put overall agreement at 95% and agreement for every unhealthy category at 92.5% or higher, with fast food at 100% (Supplementary Table S8), so the categories we report rarely contain something that does not belong. The harder question is recall, particularly for fast food: because we identify fast food from a curated chain list, independent and fast-casual quick-service outlets that the source data codes as full-service are missed, and the validation sample contained examples, such as a hot- dog stand and a fast-casual bakery-cafe sitting among full-service restaurants. We therefore likely undercount fast food, which would if anything understate the unhealthy retail presence, and we treat the fast-food count as a lower bound; convenience stores and off-premise alcohol, which dominate the unhealthy set and are coded more reliably, carry the income gradient. Second, we characterise the retail environment, not diet or health. The link from outlet density to individual behaviour is itself contested (Caspi et al., 2012; Sturm and Hattori, 2015), and we make no causal claim about health outcomes. Third, the exposure is area-level and uses residential population as the denominator, which does not capture where residents actually shop, work, or travel, a known limitation of place-based food-environment measures; we also apply a single set of American Community Survey 2018-2022 tract estimates to every year from 2016 to 2025, so the denominators and covariates do not track within-decade demographic change. Fourth, the two earliest annual files differ in classification composition in a way consistent with a one-time coverage or coding change rather than real change on the ground; we addressed this by repeating the flow models on a 2018-onward window, with essentially unchanged results (Supplementary Table S7). Fifth, Rhode Island is a single, small, densely settled state with essentially no rural tracts and its own alcohol-licensing regime; the magnitudes we report, and the alcohol result in particular, may not transfer to settings with different retail geographies or licensing rules. The compensating strength is completeness: the panel is an exhaustive census of one state’s establishments with a true longitudinal link rather than a sample of many places. Sixth, the openings models are associational; we cannot rule out that unmeasured neighbourhood characteristics drive both lower income and a higher rate of new openings, and because that higher rate is not specific to unhealthy retail (Section 3.6), we do not identify why commercial entry as a whole favours lower-income neighbourhoods, only that the unhealthy gradient is reproduced along with it. Finally, HOLC grades exist only for the 92 urbanised tracts, so the redlining results apply to that subset.

## 5. Conclusion

Across a decade and the entire state, the unhealthy retail environment was concentrated in lower-income Rhode Island neighbourhoods, and that concentration was stable and robust to how it was measured. The gradient was reproduced through entry: new outlets arrived more often in lower-income neighbourhoods, while the outlets already present left at a similar rate regardless of neighbourhood income, so exit did not explain the pattern. This entry was not unhealthy- specific but reflected a broad tilt of commercial activity toward lower-income neighbourhoods. Efforts to reshape the retail environment in these neighbourhoods will therefore have more purchase on the flow of new openings than on the existing stock, provided they are defined by the convenience and off-premise alcohol formats that make up most of the standing burden rather than by a blunt limit on entry that would also curb the healthier options those neighbourhoods gain. Characterising how such environments are produced, and monitoring them as they change, is a prerequisite for acting on them.

## Declarations

### Ethics

This study analysed commercial business records and publicly available area-level census data and did not involve human participants or identifiable personal information; institutional review board approval was therefore not required.

## Data availability

The business establishment records were obtained under licence from Data Axle USA and cannot be redistributed by the authors. The American Community Survey estimates and TIGER/Line boundaries are publicly available from the United States Census Bureau, and the Historic Redlining Indicator is available from the Inter-university Consortium for Political and Social Research (https://doi.org/10.3886/E141121V3).

## Code availability

The analysis code will be made available in a public repository upon acceptance.

## Acknowledgements

The authors acknowledge Data Axle USA for provision of business establishment records under licence, and the U.S. Census Bureau for the American Community Survey and TIGER/Line data used in this study.

## References

Allison, P.D., 1982. Discrete-time methods for the analysis of event histories. Sociological Methodology 13, 61–98.

Besag, J., York, J., Mollié, A., 1991. Bayesian image restoration, with two applications in spatial statistics. Annals of the Institute of Statistical Mathematics 43 (1), 1–20.

Cameron, A.C., Miller, D.L., 2015. A practitioner’s guide to cluster-robust inference. Journal of Human Resources 50 (2), 317–372.

Campbell, C.A., Hahn, R.A., Elder, R., Brewer, R., Chattopadhyay, S., Fielding, J., Naimi, T.S., Toomey, T., Lawrence, B., Middleton, J.C., Task Force on Community Preventive Services, 2009. The effectiveness of limiting alcohol outlet density as a means of reducing excessive alcohol consumption and alcohol-related harms. American Journal of Preventive Medicine 37 (6), 556–569.

Caspi, C.E., Sorensen, G., Subramanian, S.V., Kawachi, I., 2012. The local food environment and diet: a systematic review. Health & Place 18 (5), 1172–1187.

Cooksey-Stowers, K., Schwartz, M.B., Brownell, K.D., 2017. Food swamps predict obesity rates better than food deserts in the United States. International Journal of Environmental Research and Public Health 14 (11), 1366.

Han, E., Powell, L.M., Zenk, S.N., Rimkus, L., Ohri-Vachaspati, P., Chaloupka, F.J., 2012. Classification bias in commercial business lists for retail food stores in the U.S. International Journal of Behavioral Nutrition and Physical Activity 9, 46.

Lee, E.K., Donley, G., Ciesielski, T.H., Gill, I., Yamoah, O., Roche, A., Martinez, R., Freedman, D.A., 2022. Health outcomes in redlined versus non-redlined neighbourhoods: a systematic review and meta-analysis. Social Science & Medicine 294, 114696.

Li, M., Yuan, F., 2022. Historical redlining and food environments: a study of 102 urban areas in the United States. Health & Place 75, 102775.

Liese, A.D., Colabianchi, N., Lamichhane, A.P., Barnes, T.L., Hibbert, J.D., Porter, D.E., Nichols, M.D., Lawson, A.B., 2010. Validation of 3 food outlet databases: completeness and geospatial accuracy in rural and urban food environments. American Journal of Epidemiology 172 (11), 1324–1333.

Lovasi, G.S., Boise, S., Jogi, S., Hurvitz, P.M., Rundle, A.G., Diez, J., Hirsch, J.A., Fitzpatrick, A., Biggs, M.L., Siscovick, D.S., 2023. Time-varying food retail and incident disease in the Cardiovascular Health Study. American Journal of Preventive Medicine 64 (6), 877–887.

Meier, H.C.S., Mitchell, B.C., 2023. Historic Redlining Indicator for 2000, 2010, and 2020 US Census Tracts. Inter-university Consortium for Political and Social Research [distributor], Ann Arbor, MI. 10.3886/E141121V3

Moore, L.V., Diez Roux, A.V., 2006. Associations of neighbourhood characteristics with the location and type of food stores. American Journal of Public Health 96 (2), 325–331.

Rhode Island Department of Business Regulation, 2023. The Rhode Island liquor control law (Title 3) and the three-tier system. State of Rhode Island, Providence, RI.

Riebler, A., Sørbye, S.H., Simpson, D., Rue, H., 2016. An intuitive Bayesian spatial model for disease mapping that accounts for scaling. Statistical Methods in Medical Research 25 (4), 1145–1165.

Rue, H., Martino, S., Chopin, N., 2009. Approximate Bayesian inference for latent Gaussian models by using integrated nested Laplace approximations. Journal of the Royal Statistical Society: Series B (Statistical Methodology) 71 (2), 319–392.

Shannon, J., 2021. Dollar stores, retailer redlining, and the metropolitan geographies of precarious consumption. Annals of the American Association of Geographers 111 (4), 1200–1218.

Sturm, R., Hattori, A., 2015. Diet and obesity in Los Angeles County 2007-2012: is there a measurable effect of the 2008 “Fast-Food Ban”? Social Science & Medicine 133, 205–211.

Wagstaff, A., Paci, P., van Doorslaer, E., 1991. On the measurement of inequalities in health. Social Science & Medicine 33 (5), 545–557.

Zhang, X., Hatcher, B., Clarkson, L., Holt, J., Bagchi, S., Kanny, D., Brewer, R.D., 2015. Changes in density of on-premises alcohol outlets and impact on violent crime, Atlanta, Georgia, 1997-2007. Preventing Chronic Disease 12, 140317.

