## Supplementary Materials for "How new openings sustain the income gradient in unhealthy retail: evidence from a statewide establishment panel, Rhode Island, 2016-2025"

### **Supplementary Material**

#### **S1. Establishment classification and counts**

Establishments were classified from NAICS (authoritative) with SIC used to resolve missing or ambiguous codes. The unhealthy set comprised liquor stores, drinking places, convenience stores, and fast food; the healthy set comprised supermarkets and grocery stores and produce markets; coffee, donut, and ice cream outlets and full-service restaurants were treated as neutral; tobacco-only retailers were excluded. Two NAICS codes were reconciled across the 2022 to 2023 vintage boundary: liquor stores (445310 to 445320) and convenience stores (445120 to 445131). The 2019 annual file duplicated an adjacent year and was dropped, leaving nine cross-sections. Classification was checked against a stratified random sample of 360 establishment-records, 40 in each of the nine assigned categories, reviewed by hand against business name and full NAICS and SIC descriptions; overall agreement was 95.0% (Supplementary Table S8).

Between 2016 and 2017 the recorded convenience-store count rises by 56 percent (299 to 467) and the bar count falls by 55 percent (220 to 98), while liquor stores and fast food are essentially flat. The establishment-identifier flows in Supplementary Table S6 show that these are reclassifications of businesses already present rather than genuine entries or exits: only 20 cohort establishments first appear in 2017. We therefore treat the 2016 to 2017 change as a coverage and coding shift between the two earliest files, and report a 2018-onward sensitivity analysis for the flow models (Supplementary Table S7).

**Supplementary Table S1.** Establishment counts by category and year.

| **Category** | **2016** | **2017** | **2018** | **2020** | **2021** | **2022** | **2023** | **2024** | **2025** |
| --- | --- | --- | --- | --- | --- | --- | --- | --- | --- |
| convenience | 299 | 467 | 427 | 449 | 409 | 400 | 390 | 413 | 445 |
| drinking_places | 220 | 98 | 106 | 141 | 131 | 133 | 174 | 183 | 188 |
| fast_food | 313 | 311 | 292 | 291 | 258 | 259 | 272 | 275 | 264 |
| full_service | 2143 | 2511 | 2316 | 2354 | 2231 | 2250 | 2336 | 2564 | 2654 |
| liquor_stores | 232 | 223 | 223 | 225 | 220 | 209 | 212 | 230 | 225 |
| other | 45592 | 43766 | 44395 | 49920 | 49166 | 49018 | 51433 | 53981 | 56813 |
| produce_markets | 15 | 12 | 10 | 10 | 11 | 12 | 14 | 15 | 16 |
| snack_coffee | 606 | 420 | 431 | 431 | 395 | 401 | 415 | 431 | 440 |
| supermarket_grocery | 256 | 232 | 222 | 258 | 273 | 276 | 283 | 290 | 301 |

Unhealthy outlets are the sum of convenience, drinking_places, fast_food, and liquor_stores; healthy outlets are the sum of produce_markets and supermarket_grocery. The counts above total 481,526 establishment-years; the 480,923 reported in the main text and used in the models exclude 603 establishment-years whose geocoordinates fell outside all Rhode Island census tracts (coastal or out-of-state points).

#### **S2. Concentration index by year**

The population-weighted health concentration index ranks tracts by the socioeconomic variable and summarises how unequally unhealthy outlets per capita are distributed across that ranking. Negative values indicate concentration among lower-ranked (here, lower-income or lower non-Hispanic White share) tracts. Confidence intervals are from a nonparametric bootstrap.

**Supplementary Table S2.** Concentration index for unhealthy outlets per capita, by year.

| **Ranked by** | **Year** | **CI** | **Lo** | **Hi** |
| --- | --- | --- | --- | --- |
| income | 2016 | -0.1647 | -0.215 | -0.1094 |
| income | 2017 | -0.166 | -0.2114 | -0.1109 |
| income | 2018 | -0.1639 | -0.2131 | -0.1088 |
| income | 2020 | -0.1592 | -0.2117 | -0.1028 |
| income | 2021 | -0.1596 | -0.2122 | -0.1034 |
| income | 2022 | -0.1524 | -0.2054 | -0.0945 |
| income | 2023 | -0.1593 | -0.213 | -0.1021 |
| income | 2024 | -0.1659 | -0.2191 | -0.1074 |
| income | 2025 | -0.1711 | -0.2207 | -0.1143 |
| pct_white | 2016 | -0.0913 | -0.1425 | -0.0403 |
| pct_white | 2017 | -0.1065 | -0.1535 | -0.0587 |
| pct_white | 2018 | -0.0963 | -0.1436 | -0.0479 |
| pct_white | 2020 | -0.1017 | -0.1501 | -0.052 |
| pct_white | 2021 | -0.0982 | -0.1466 | -0.0476 |
| pct_white | 2022 | -0.0853 | -0.1358 | -0.033 |
| pct_white | 2023 | -0.0956 | -0.1496 | -0.0417 |
| pct_white | 2024 | -0.0904 | -0.1465 | -0.0354 |
| pct_white | 2025 | -0.096 | -0.1495 | -0.0416 |

The income concentration index is consistently stronger (more negative) than the non-Hispanic White index, and both are stable across the decade.

#### **S3. Stability of the income gradient by year**

The per-capita income model was refitted in each year. For the HOLC-graded subset, the same model was refitted with the standardised redlining indicator added.

**Supplementary Table S3.** Income and redlining rate ratios by year.

| **Measure** | **Year** | **IRR** | **Lo** | **Hi** | **p** |
| --- | --- | --- | --- | --- | --- |
| income IRR (full state) | 2016 | 0.716 | 0.632 | 0.811 | 1.5e-07 |
| income IRR (full state) | 2017 | 0.709 | 0.631 | 0.796 | 5.71e-09 |
| income IRR (full state) | 2018 | 0.697 | 0.619 | 0.785 | 2.76e-09 |
| income IRR (full state) | 2020 | 0.719 | 0.637 | 0.81 | 6.19e-08 |
| income IRR (full state) | 2021 | 0.713 | 0.631 | 0.806 | 5.35e-08 |
| income IRR (full state) | 2022 | 0.723 | 0.638 | 0.819 | 3.27e-07 |
| income IRR (full state) | 2023 | 0.727 | 0.642 | 0.824 | 6.29e-07 |
| income IRR (full state) | 2024 | 0.709 | 0.626 | 0.803 | 5.87e-08 |
| income IRR (full state) | 2025 | 0.705 | 0.622 | 0.798 | 3.64e-08 |
| redlining HRI IRR (HOLC subset) | 2016 | 1.16 | 1.007 | 1.336 | 0.0392 |
| redlining HRI IRR (HOLC subset) | 2017 | 1.129 | 0.991 | 1.287 | 0.0689 |
| redlining HRI IRR (HOLC subset) | 2018 | 1.132 | 0.987 | 1.299 | 0.0769 |
| redlining HRI IRR (HOLC subset) | 2020 | 1.129 | 0.984 | 1.296 | 0.0844 |
| redlining HRI IRR (HOLC subset) | 2021 | 1.138 | 0.989 | 1.309 | 0.0712 |
| redlining HRI IRR (HOLC subset) | 2022 | 1.11 | 0.963 | 1.281 | 0.151 |
| redlining HRI IRR (HOLC subset) | 2023 | 1.123 | 0.972 | 1.297 | 0.115 |
| redlining HRI IRR (HOLC subset) | 2024 | 1.138 | 0.982 | 1.319 | 0.0855 |
| redlining HRI IRR (HOLC subset) | 2025 | 1.158 | 1 | 1.342 | 0.0501 |

The income gradient is stable year to year. The redlining association in the graded subset is positive but mostly does not reach significance once the model is fit year by year.

#### **S4. Redlining models (secondary)**

These models are restricted to the 92 HOLC-graded tracts and are reported as a secondary analysis. The standardised Historic Redlining Indicator (hri_z) is entered alone and then with standardised income (inc_z) and log population density (logpd_z).

**Supplementary Table S4.** Negative binomial models of unhealthy outlets per capita with the Historic Redlining Indicator, most recent year.

| **Model** | **Term** | **Estimate** | **Lo** | **Hi** | **p** |
| --- | --- | --- | --- | --- | --- |
| HRI alone | hri_z | 1.25 | 1.085 | 1.44 | 0.00202 |
| HRI + income + urbanicity | hri_z | 1.159 | 1 | 1.344 | 0.0501 |
| HRI + income + urbanicity | inc_z | 0.821 | 0.702 | 0.961 | 0.0142 |
| HRI + income + urbanicity | logpd_z | 0.857 | 0.751 | 0.977 | 0.0213 |

The unadjusted redlining association attenuates to the margin of significance after adjustment for present-day income and density, while income remains associated. Closure and opening models within the graded subset appear in Table 4 of the main text.

#### **S5. Bayesian spatial model specification**

The BYM2 model (Besag et al., 1991; Riebler et al., 2016) was fitted with R-INLA (Rue et al., 2009) as a Poisson rate model with tract resident population as the expected count. Fixed effects were standardised income and standardised log population density. The random effect was a BYM2 term on a queen-contiguity tract adjacency, with the spatial component scaled (scale.model) and penalised-complexity priors: a PC prior on the precision with parameters (1, 0.01) and a PC prior on the mixing parameter phi with parameters (0.5, 0.5). The fitted mixing parameter phi was 0.061. Because the penalised-complexity prior placed on this parameter, with parameters (0.5, 0.5), favours smaller structured shares, we read this as evidence against strong spatially structured residual variation once income and density are included, rather than as a precise estimate of the structured share; we do not interpret it as showing that the covariates fully absorb the spatial pattern. The same BYM2 specification applied to period-aggregated unhealthy openings (Poisson, with tract resident population as the expected count) likewise showed the opening rate falling with income (rate ratio 0.739, 95% credible interval 0.622 to 0.879), with a spatial mixing parameter phi of 0.108, essentially reproducing the non-spatial openings estimate of 0.734.

#### **S6. Establishment flows by year**

The modal-unhealthy cohort comprises 1,528 establishments. The table below counts, in each observed wave, the number whose establishment identifier first appears (new appearances), the number last seen in that wave, the resulting closures (a last-seen wave before the end of the panel), and the number active. New appearances in the first observed wave (2016) are establishments already in operation at the panel’s start, not openings.

Supplementary Table S6. Establishment-identifier flows by year, modal-unhealthy cohort.

| **Year** | **New appearances** | **Last seen** | **Closures** | **Active** |
| --- | --- | --- | --- | --- |
| 2016 | 1,081 | 30 | 30 | 1,081 |
| 2017 | 20 | 134 | 134 | 1,071 |
| 2018 | 111 | 68 | 68 | 1,048 |
| 2020 | 111 | 107 | 107 | 1,091 |
| 2021 | 31 | 24 | 24 | 1,015 |
| 2022 | 11 | 33 | 33 | 1,002 |
| 2023 | 74 | 20 | 20 | 1,043 |
| 2024 | 50 | 43 | 43 | 1,073 |
| 2025 | 39 | 1,069 | 0 | 1,069 |

Openings are establishments first appearing after 2016 (the sum of new appearances in 2017 and later, 447). Closures are establishments last seen before 2025 (459). The remaining 1,069 are present in the final wave. Of the 20 establishments first appearing in 2017, 12 are convenience stores, 5 fast food, 2 liquor stores, and 1 a bar; this is the same period over which the recorded convenience count rises by 56 percent (Supplementary Table S1), confirming that the rise reflects reclassification of businesses already in operation rather than new entry. Because the 2019 file is absent, the 68 establishments last seen in 2018 cannot be separated from possible 2019 closures; person-periods treat 2018 and 2020 as adjacent waves.

#### **S7. Baseline sensitivity of the flow models**

To check that the openings and closures results do not depend on the anomalous 2016 to 2017 transition, we repeated both flow models on a 2018-onward window, treating 2016 and 2017 as a settling-in period. The full-panel models use 9,475 person-periods, 447 openings, and 459 closures; the 2018-onward models use 7,327 person-periods, 427 openings, and 295 closures. Estimates are rate ratios (openings) or hazard ratios (closures) per standard deviation, with 95% intervals and p values.

**Supplementary Table S7.** Flow models under the full panel and the 2018-onward window.

| **Analysis** | **Sample** | **Term** | **Full panel (2016-2025)** | **2018-onward window** |
| --- | --- | --- | --- | --- |
| Closure hazard | full state | income | 1.105 (0.986-1.239), p=0.086 | 1.126 (0.969-1.309), p=0.122 |
| Closure hazard | full state | log pop density | 1.206 (1.072-1.356), p=0.002 | 1.229 (1.053-1.433), p=0.009 |
| Openings | full state | income | 0.734 (0.595-0.906), p=0.004 | 0.730 (0.586-0.910), p=0.005 |
| Openings | full state | log pop density | 1.064 (0.890-1.273), p=0.50 | 1.034 (0.861-1.243), p=0.72 |
| Closure hazard | HOLC subset | redlining (HRI) | 1.172 (1.005-1.367), p=0.043 | 1.259 (1.033-1.534), p=0.023 |
| Openings | HOLC subset | redlining (HRI) | 1.303 (0.954-1.780), p=0.096 | 1.330 (0.952-1.860), p=0.095 |

The income gradient in openings is essentially identical under the two settings (rate ratio 0.734 versus 0.730), and the closure hazard remains unrelated to income under both (hazard ratio 1.105 versus 1.126). The descriptive contrast between the lowest- and highest-income tertiles in openings per resident is likewise preserved (roughly 2.2-fold on the full panel and similar on the 2018-onward window; the harmonised tract-level tertile rates for the full panel appear in Table 5). The closure count is roughly a third smaller on the restricted window, because the elevated 2016 and 2017 last-seen records are excluded, but the even-handedness of closures across income does not change. One asymmetry is worth noting: an establishment first observed in 2018 is counted as an opening on this window, while one first observed in 2016 is treated as pre-existing stock. This rests on observing the later entrant’s absence from the 2016 and 2017 files, and because the openings income gradient is essentially identical whether the window starts in 2016 or 2018, it is not sensitive to where that boundary is drawn.

#### **S8. Classification validation**

A stratified random sample of 40 establishment-records per assigned category (360 in total across the nine categories) was reviewed by hand against business name and full NAICS and SIC descriptions. Agreement is the share of records whose assigned category was judged correct, that is, precision. Confidence intervals are Wilson score intervals.

**Supplementary Table S8.** Hand validation of the classification: agreement (precision) by assigned category.

| **Assigned category** | **Reviewed** | **Correct** | **Agreement** | **95% CI** |
| --- | --- | --- | --- | --- |
| Convenience | 40 | 37 | 0.925 | 0.801-0.974 |
| Drinking places | 40 | 39 | 0.975 | 0.871-0.996 |
| Fast food | 40 | 40 | 1.000 | 0.912-1.000 |
| Liquor stores | 40 | 39 | 0.975 | 0.871-0.996 |
| Supermarket and grocery | 40 | 36 | 0.900 | 0.769-0.960 |
| Produce markets | 40 | 38 | 0.950 | 0.835-0.986 |
| Snack and coffee (neutral) | 40 | 36 | 0.900 | 0.769-0.960 |
| Full-service restaurants (neutral) | 40 | 37 | 0.925 | 0.801-0.974 |
| Other (excluded) | 40 | 40 | 1.000 | 0.912-1.000 |
| Overall | 360 | 342 | 0.950 | 0.922-0.968 |

Disagreements were predominantly source-data coding errors, in which a business carrying a food-retail industry code was not in fact a food outlet (for example a car wash, an auto-service station, a nonprofit, or a facility-management firm coded under a grocery or liquor code). A smaller number reflected category edges that do not affect the healthy-versus-unhealthy split: a few pizza outlets and a caterer fell into the neutral snack-and-coffee group rather than fast food. This procedure measures precision. Recall for fast food is addressed in the Limitations: the curated chain list does not capture independent or fast-casual quick-service outlets that the source codes as full-service, so the fast-food count is best read as a lower bound.

#### **S9. Net flow and stock accounting by income tertile**

To show how a stable standing gradient coexists with turnover, the table below gives, for each tract income tertile over 2016 to 2025, the counts of openings and closures, the resulting net flow, and the per-capita unhealthy stock in the first and last observed waves. Tertiles are tract-level; the panel covers the 1,526 of 1,528 cohort establishments whose tract had valid covariates, and the flow totals reconcile with Supplementary Table S6 (447 openings, 459 closures).

**Supplementary Table S9.** Openings, closures, net flow, and per-capita stock by tract income tertile.

| **Income tertile** | **Openings** | **Closures** | **Net** | **Stock per 1,000, 2016** | **Stock per 1,000, 2025** |
| --- | --- | --- | --- | --- | --- |
| Low | 204 | 188 | +16 | 1.284 | 1.330 |
| Middle | 140 | 155 | -15 | 1.039 | 0.996 |
| High | 103 | 116 | -13 | 0.676 | 0.643 |

The lowest-income tertile runs a small net gain in unhealthy outlets over the decade while the middle and highest tertiles run net losses, yet the per-capita stock ratio between the lowest- and highest-income tertiles moves only from about 1.9 to about 2.1. The stability of the gradient is thus a balance of flows: an even closure rate applied to the larger low-income stock removes more outlets there in absolute terms, offsetting the higher rate of openings.

#### **S10. Category-specific gradients and the effect of excluding bars**

The unhealthy set pools four outlet types. The table below reports, for each type, the cross-sectional per-capita income rate ratio (most recent year) and the income rate ratio for openings, each per standard deviation of higher income. All four types share the same cross-sectional gradient; the openings gradients run in the same direction for all four but reach significance only for convenience stores, reflecting the smaller opening counts within any single type.

**Supplementary Table S10a.** Category-specific income gradients (rate ratio per SD income, 95% interval).

| **Category** | **Stock rate ratio (most recent year)** | **Openings rate ratio** |
| --- | --- | --- |
| Liquor stores | 0.786 (0.637-0.969) | 0.769 (0.538-1.099) |
| Bars (drinking places) | 0.660 (0.451-0.968) | 0.719 (0.474-1.091) |
| Convenience stores | 0.679 (0.575-0.801) | 0.738 (0.571-0.953) |
| Fast food | 0.679 (0.522-0.884) | 0.785 (0.533-1.158) |

The recorded number of bars is unstable across the panel (220, 98, 106, 141, 131, 133, 174, 183, and 188 in 2016 through 2025; Supplementary Table S1), consistent with drifting classification rather than real change. Removing bars from the unhealthy set leaves the flow result essentially unchanged, confirming that convenience stores and off-premise alcohol carry the gradient.

**Supplementary Table S10b.** Flow models with and without bars (income effect per SD).

| **Unhealthy set** | **Cohort** | **Openings** | **Closures** | **Openings RR (income)** | **Closure HR (income)** |
| --- | --- | --- | --- | --- | --- |
| All four types | 1,528 | 447 | 459 | 0.734 (p=0.004) | 1.105 (p=0.086) |
| Excluding bars | 1,222 | 257 | 358 | 0.757 (p=0.004) | 1.113 (p=0.11) |
